# Unraveling the role of non-coding rare variants in epilepsy

**DOI:** 10.1101/2022.12.13.22283363

**Authors:** Alexandre Girard, Claudia Moreau, Jacques Michaud, Berge Minassian, Patrick Cossette, Simon L. Girard

**Author notes:** **Contact info** Simon Girard, PhD, Université du Québec à Chicoutimi, Département des sciences fondamentales, Office P4-2130, 555, boulevard de l’Université, Chicoutimi (Québec) G7H 2B1, 1 (toll free).

## Abstract

**Importance:** Despite the use of very large cohorts, the discovery of new variants has leveled off in recent years in epilepsy studies and consequently, most of the heritability is still unexplained. Rare non-coding variants have been largely ignored in studies on epilepsy, although non-coding single nucleotide variants can have a significant impact on gene expression.

**Objective:** To determine if rare non-coding deleterious variants are associated with epilepsy.

**Design:** This is a case-control study made possible by the CENet cohort.

**Setting:** This was initially a multicenter study (families and trios), although the sequencing was processed at the same facility and for the present case-control study, only unrelated individuals were drawn.

**Participants:** Patients used in this study are affected either by genetic generalized epilepsy (GGE), non-acquired focal epilepsy (NAFE) or are called ‘mixed’ (phenotype that differs from other affected relatives). Controls are unaffected parents from developmental and epileptic encephalopathy trios.

**Main Outcomes and Measures:** To assess the functional impact of non-coding variants, ExPecto, a deep learning algorithm was used. A binomial logistic regression was performed to compare the burden of rare non-coding deleterious variants between cases and controls.

**Results:** We had access to WGS from 123 GGE, 112 NAFE and 12 mixed for a total of 247 patients, as well as 377 controls. Rare non-coding highly deleterious variants were associated with GGE (OR 2.74; 95% CI 1.20-6.22), but not with NAFE (OR 0.85; 95% CI 0.27-2.67) or all epilepsy cases (OR 1.54; 95% CI 0.77-3.11) when compared with controls.

**Conclusion and Relevance:** In this study we showed that rare non-coding deleterious variants are associated with epilepsy, specifically with GGE. Larger WGS epilepsy cohort will be needed to investigate those effects at a greater resolution. Nevertheless, we demonstrated the importance of studying non-coding regions in epilepsy, a disease where new discoveries are scarce, and a high proportion of the heritability is yet to be explained.

**Key points:** *Question:* Are non-coding single nucleotide variants (SNV) associated with epilepsy?

*Findings:* In this study we showed that patients with generalized genetic epilepsy (GGE) had significantly more rare non-coding deleterious variants than controls and non-acquired focal epilepsy (NAFE) patients. The study included 247 epilepsy patients and 377 controls who were sequenced for the whole genome.

*Meaning:* Rare non-coding SNV are associated with epilepsy, more specifically with GGE.

## Introduction

Epilepsy is a neurological disorder characterized by epileptic seizures and spontaneous episodes of abnormal neuronal activity^1^. Approximately 3% of all individuals will be affected during their lifetime ^2^. New variant discoveries are rarer in recent epilepsy studies. Only large cohorts composed of tens of thousands of individuals had some success^3–6^. Those studies mainly focused on common or coding variants^5,6^. Indeed, a large proportion of the heritability of the disease is still unexplained^5^. The overwhelming majority of studies in epilepsy use either genotyping or exome sequencing to investigate the genetic causes of the disease. Consequently, little is known concerning the implication of non-coding regions in the etiology of the disease^3–6^. However, these regions were shown to have an important impact on the phenotype of an individual as they affect the expression of neighboring genes^7^.

Studying expression quantitative trait loci (eQTL) in neurological disease is a notable challenge, mainly because eQTLs effects are tissue specific and brain tissues are only available post-mortem. Nevertheless, progress in deep learning now allows us to predict the functional effects of variants from sequencing data alone^8–10^. We aimed to characterize the role of rare non-coding variants in epilepsy based on their functional effect in brain tissues using the most cited deep learning algorithm, ExPecto^8^. We used whole genome sequencing (WGS) data from the Canadian Epilepsy Network (CENet) cohort^11^ to investigate deleterious rare functional variants in epileptic patients.

## Methods

### Subjects

We used WGS from 247 unrelated patients, 123 who have Genetic Generalized Epilepsy (GGE), 112 who have Non-Acquired Focal Epilepsy (NAFE) and 12 patients who have a ‘Mixed’ phenotype (a different subtype of epilepsy than their affected relatives). Controls are 377 healthy parents having at least one child affected by Developmental Epileptic Encephalopathy (DEE) (Table 1). Patients were recruited in CHUM Research Center in Montreal and controls in the hospital for Sick Children in Toronto and CHU Ste-Justine in Montreal. All samples were sequenced at Genome Quebec Innovation Center in Montreal. See eMethods for more details on phenotyping and sequencing.

**Table 1.** Number of individuals for each phenotype

| **Phenotype** | **Male** | **Female** | **Total** |
| --- | --- | --- | --- |
| Controls | 190 | 187 | 377 |
| All Cases | 112 | 135 | 247 |
| GGE | 52 | 71 | 123 |
| NAFE | 50 | 62 | 112 |
| Mixed | 10 | 2 | 12 |

### Analyses

Single nucleotide variants (SNVs) were filtered based on their minor allele frequency (maf), only rare variants (maf <0.01) both in our cohort and in gnomAD^12^ were kept. We applied ExPecto^8^ on rare variants for three tissues related to epilepsy: hippocampus, amygdala and brain cortex for which we calculated the median^13^. The accuracy of ExPecto predictions was validated using known eQTLs from the GTEx V6p release^14^ (eMethods and eFigure 1). Gene expression change calculated by ExPecto was used to compute a constraint violation score (CVS) in accordance with the methods described in^8^. The CVS quantifies how deleterious the gene expression change is: the higher the score, the more deleterious the variant.

We then compared the proportion of patients and controls who had at least one rare variant at different CVS thresholds using a logistic regression analysis with the python package statsmodels v0.12.2 to compute odds ratios (OR). Sex was used as a covariate as well as a two dimensions UMAP (umap-learn v0.5.1) based on the first 5 principal components (plink v2.0) (eMethods and eFigure 2). We repeated the analysis to compare GGE with controls, NAFE with controls and GGE with NAFE (mixed patients were removed from these analyses). Variants included in the final analysis are available in the eTable 1.

## Results

Our analyses showed no significant difference when comparing cases and controls (Figure 1A). Nevertheless, the OR tends to increase with the CVS threshold and reaches a peak for variants with a CVS above 40 (OR 1.54; 95% CI 0.77-3.11). However, there is a significant difference between GGE and controls for CVS above 40 (OR 2.74; 95% CI 1.20-6.22) (Figure 1B). On the other hand, NAFE and controls show no significant difference, meaning that the trend observed when comparing all cases and controls was solely driven by the GGE (Figure 1C). Finally, GGE and NAFE have a significantly different burden for CVS thresholds of]20, 30] (OR 2.47; 95% CI 1.10-5.52) and above 40 (OR 3.19; 95% CI 1.002-10.13) (Figure 1D).

**Figure 1.**
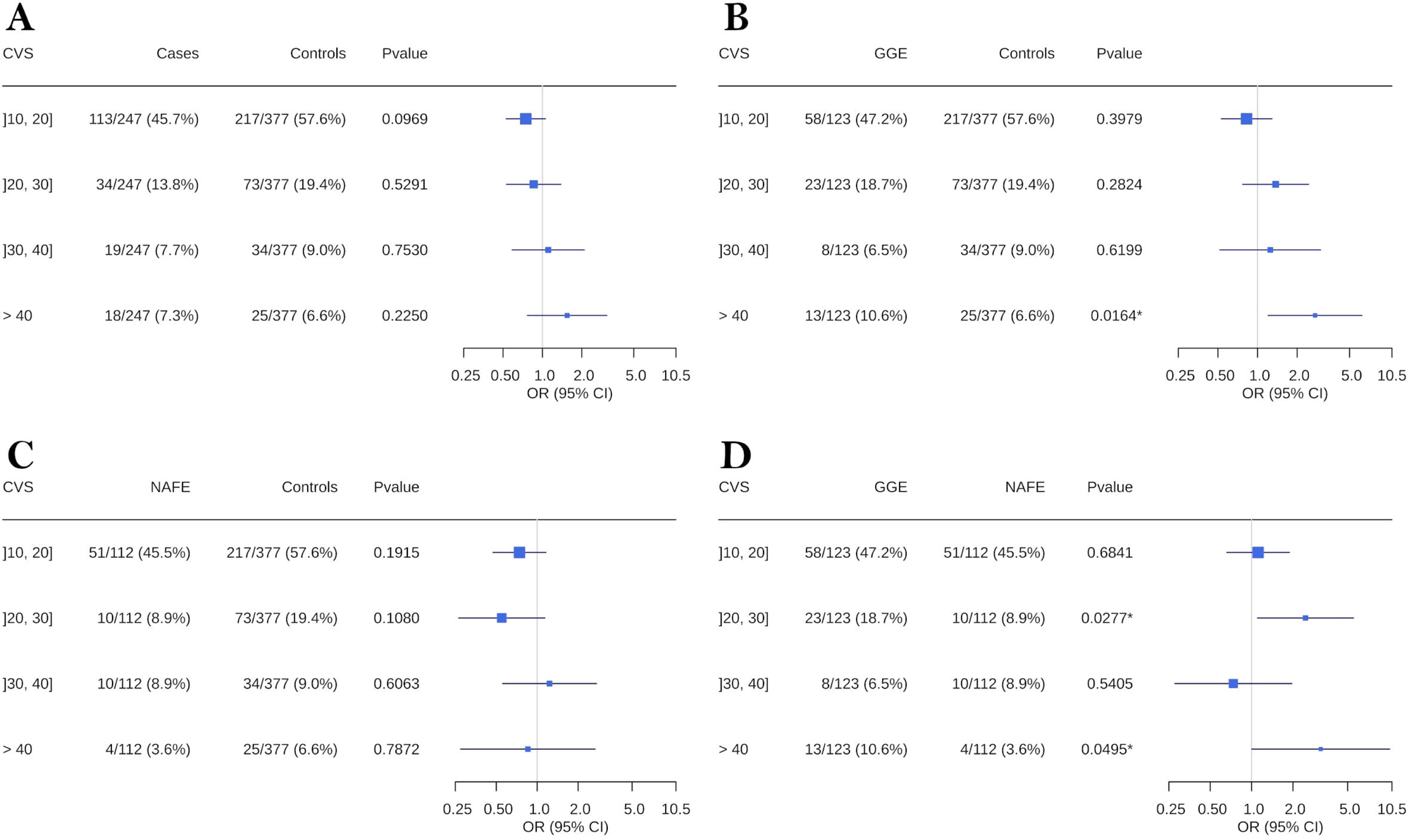
Burden of variants for different CVS thresholds across epilepsy phenotypes. Odds ratios and p-value were calculated using a binomial logistic regression for variants of different CVS thresholds. Lines represent 95% confidence intervals. Comparisons were made for cases and controls (A), GGE and controls (B), NAFE and controls (C) and GGE and NAFE (D).

## Discussion

We demonstrated that more deleterious non-coding rare variants result in higher OR in epilepsy. A trend observed when comparing all cases or only GGE with controls. However, it is not the case for NAFE who are known to have a smaller genetic burden^3^. A recent hypothesis suggests that this could be explained by a greater heterogeneity in NAFE patients who should be further divided into subphenotypes^6^. Additionally, we showed that GGE have significantly more non-coding highly deleterious variants than controls. Once again, our findings demonstrate the importance of separating GGE and NAFE when studying epilepsy, which is also supported by most associated genes being specific to subphenotypes^11,15^. We are the first to highlight the impact of rare non-coding SNV on a genome-wide scale in epilepsy. This discovery reveals the importance of studying non-coding regions which may explain a part of the missing heritability in epilepsy^5^.

### Limitations

The study has two main limitations. First, the use of deep learning, as useful as it is, has the limitation of being predictions, not observations. Nonetheless, we validated those predictions were accurate by using experimental data from GTEx (eFigure 1). Second, is our small sample size. Despite this, we were successful in identifying genome-wide effects, but we lacked power to investigate those effects at a gene or variant level resolution.

## Conclusion

This study demonstrates the importance of non-coding regions in the etiology of epilepsy. The effect was specific for GGE, whereas NAFE showed no significant difference with controls. Therefore, our results indicate that the differences between those subphenotypes extends to non-coding genetic mechanisms. Larger WGS cohorts will be needed to deepen our understanding of the role of non-coding regions in epilepsy.

## Supporting information

Supplemental Online Content

eTable 1

## Data Availability

Raw whole genome sequences of a subset of the epilepsy patients for which we have appropriate consent have been deposited in the European Genome-phenome Archive, under the accession code EGAS00001002825. The rest of the data are available upon request to the authors.

