## Supplemental Online Content for "Unraveling the role of non-coding rare variants in epilepsy"

**eMethods**

**eFigure1.** Accuracy of predictions' directionality on known GTEx v6p eQTLs.

**eFigure2.** UMAP of ethnicity for the epilepsy patients and controls.

**eReferences**

### eMethods

#### Cohort phenotyping

The epilepsy cohort is composed of extended families with affected GGE or NAFE individuals collected in CHUM Research Center in Montreal and controls were collected in CHU Ste-Justine in Montreal and the Hospital for Sick Children in Toronto as part of the Canadian Epilepsy Network (CENet)[11,16–19] . Patients were diagnosed by neurologists. The clinical epilepsy phenotype was classified according to the current classification by the International League against Epilepsy (ILAE)[20]. More specifically for NAFE, patients were at least five years of age and have experienced at least two unprovoked seizures in the six months prior to starting treatment, an MRI scan of the brain that did not demonstrate any potentially epileptogenic lesions, other than mesial temporal sclerosis. Patients with clinical and EEG characteristics meeting the 1989 ILAE syndrome definitions for GGE were included. An MRI of the brain was not required for participation. All patients were at least four years of age at the time of diagnosis. In GGE, we also included patients with Jeavons syndrome, which is an idiopathic generalized form of reflex epilepsy characterized by childhood onset, unique seizure manifestations, striking light sensitivity and possible occurrence of generalized tonic-clonic seizures. Certain cases were found with an epilepsy phenotype different from the other affected family members, hence they were marked as ‘mixed’. Controls used for this study are unaffected DEE trio parents that are from the same cohort. Only one affected GGE or NAFE patient was used per family, therefore all the individuals used in this study are unrelated.

#### Sequencing

Samples were sequenced for the whole genome at 30X coverage at Genome Quebec Innovation Center in Montreal. gDNA was cleaned using ZR-96 DNA Clean & ConcentratorTM-5 Kit (Zymo) prior to being quantified using the Quant-iTTM PicoGreen dsDNA Assay Kit (Life Technologies) and its integrity assessed on agarose gels. Libraries were generated using the TruSeq DNA PCR-Free Library Preparation Kit (Illumina) according to the manufacturer’s recommendations. Libraries were quantified using the Quant-iTTM PicoGreen dsDNA Assay Kit (Life Technologies) and the Kapa Illumina GA with Revised Primers-SYBR Fast Universal kit (Kapa Biosystems). The average size fragment was determined using a LabChip GX (PerkinElmer) instrument. The libraries were denatured in 0.05N NaOH and diluted to 8pM using HT1 buffer. The clustering was done on an Illumina cBot and the flowcell was run on a HiSeq 2 500 for 2×125 cycles (paired-end mode) using v4 chemistry and following the manufacturer’s instructions. A phiX library was used as a control and mixed with libraries at 0.01 level. The Illumina control software used was HCS 2.2.58 and the real-time analysis program used was RTA v. 1.18.64. bcl2fastq v1.8.4 was used to demultiplex samples and generate fastq reads. The filtered reads were aligned to reference Homo_sapiens assembly b37. Each readset was aligned using BWA-MEM version 0.7.10 to create a Binary Alignment Map file (.bam). Bam files were processed to gvcf files and we performed joint calling of gvcf files that were merged into a single vcf file using GATK version 3.7-0[21]. The vcf file was recalibrated and filtered following the GATK best practice guidelines.

#### Data cleaning

Cleaning was made using plink v2.0[22]. First, SNVs with a call rate below 98% were removed using ‘--geno 0.02’. Afterward, individuals with a genotype rate below 98% were excluded using ‘--mind 0.02’. Next, SNVs that did not follow the Hardy-Weinberg equilibrium were removed using ‘--hwe 0.001’. Finally, individuals with unknown biological sex were excluded. To make the principal component analysis, further cleaning was required. Only common variants were used in the PCA (maf > 0.05) and SNV that were not in linkage disequilibrium by using ‘--indep-pairwise 50 5 0.2’.

#### Validation

ExPecto was used in accordance with the method and training described in[8]. Expecto’s validation was made with eQTLs from the GTEx v6p database[13]. We used 2 parameters, the prediction’s directionality and magnitude. Directionality is defined as whether the SNV increases or decreases gene expression. Magnitude is defined as the absolute size of the effect in gene expression fold change (natural log). We determined that the magnitude above which the accuracy of the prediction’s directionality was perfect is 0.2 and we kept only variants with a median (for the cortex, hippocampus and amygdala) above this threshold (eFigure 1).

**eFigure 1.** Accuracy of predictions' directionality on known GTEx v6p eQTLs.

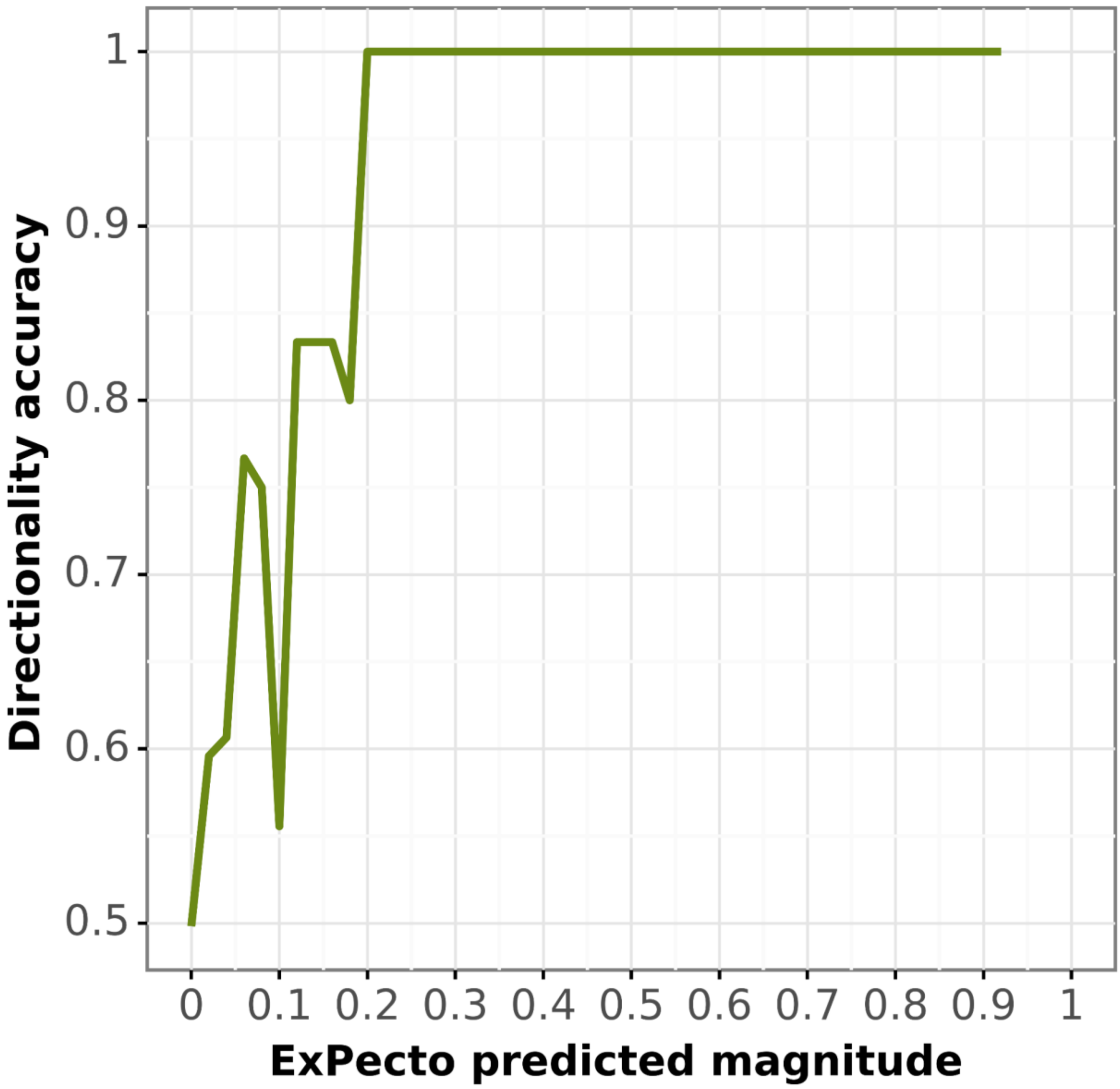


Directionality accuracy was computed according to ExPecto's predicted magnitude in natural log fold change.

**eFigure 2.** UMAP of ethnicity for the epilepsy patients and controls.

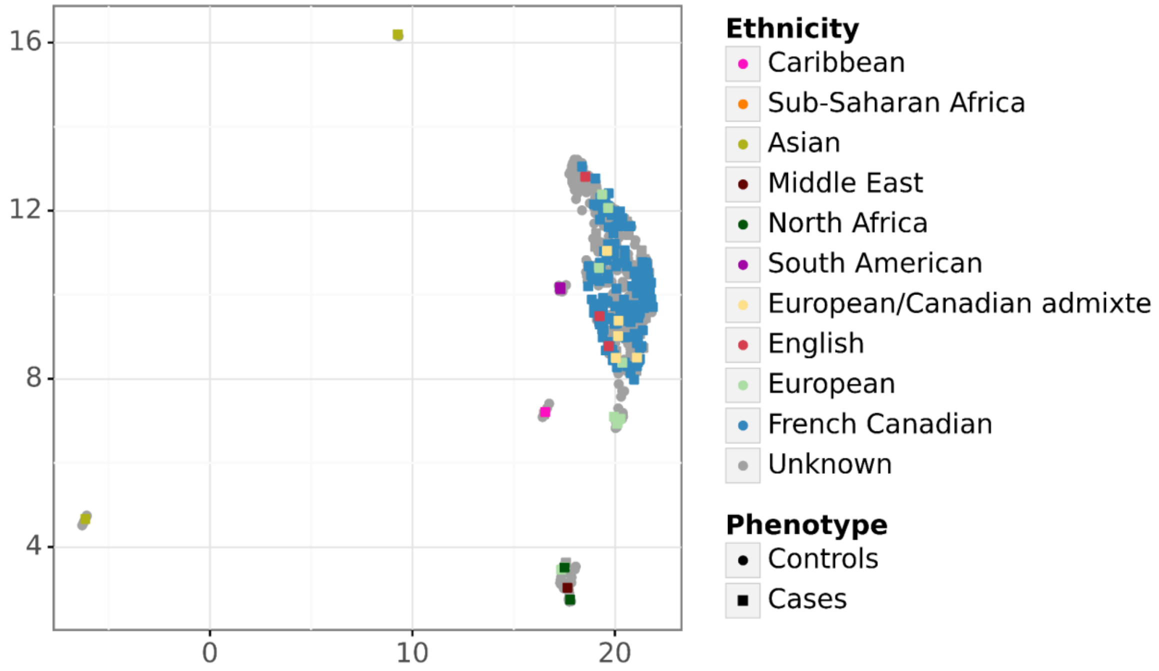


The UMAP was made with ‘umap-learn v0.5.1’ and based on the first 5 principal components.
